# Feeding Activated *Bifidobacterium infantis* EVC001 to Very Low Birth Weight Infants is Associated with Significant Reduction in Rates of Necrotizing Enterocolitis

**DOI:** 10.1101/2021.06.29.21259737

**Authors:** Joseph Tobias, Amy Olyaei, Bryan Laraway, Brian K. Jordan, Stephanie Dickinson, Lilian G. Arroyo, Elizabeth Fialkowski, Arthur Owora, Brian Scottoline

## Abstract

**Objectives:** To assess the effects of *Bifidobacterium infantis* EVC001 administration on the rate of necrotizing enterocolitis (NEC) in preterm infants in a single Level IV NICU.

**Study Design:** This was a retrospective observational analysis of 2 cohorts of VLBW infants (+/-*B. infantis* EVC001 probiotic) at OHSU from 2014 to 2020. Outcomes included NEC rates and NEC-associated mortality, including subgroup analysis of ELBW infants. Fisher’s exact test and log binomial models were used to determine differences between cohorts and risk reduction of NEC. Adjusted number needed to treat was calculated from the cohort coefficient of the model.

**Results:** In this analysis of 483 infants, the difference in rates of NEC between cohorts was statistically significant (11.0% vs. 2.7%, P = 0.0008). The EVC001-fed cohort had a 73% risk reduction of NEC compared to the No EVC001 cohort (adjusted risk ratio 0.270, 95% CI 0.094, 0.614, P = 0.0054) resulting in an adjusted number needed to treat of 12.4 (95% CI 10.0, 23.5) for *B. infantis* EVC001. There was no NEC-related mortality in the EVC001-fed cohort, yielding statistically significant differences from the No EVC001 cohort overall (0% vs. 2.7%, P = 0.0274) and the ELBW subgroup (0% vs. 5.6%, P = 0.0468).

**Conclusion(s):** *B. infantis* EVC001 feeding was associated with a significant reduction in the rate of NEC and NEC-related mortality in an observational study of 483 VLBW infants. *B. infantis* EVC001 supplementation may be considered safe and effective for reducing morbidity and mortality in the NICU.

## Introduction

Necrotizing enterocolitis (NEC) is a devastating neonatal inflammatory bowel disease that disproportionately affects preterm, very low birth weight infants (birth weight less than 1500 grams, VLBW).[1] In the United States, NEC has an incidence of 5-10% among VLBW infants and carries an overall mortality of 23.5%, although mortality reaches as high as 50% among those patients requiring surgery.[2] Survivors may experience serious long-term morbidity, including neurologic impairment and intestinal failure.[2]

NEC encompasses multiple entities with a common presentation, making the diagnosis and determination of pathobiology difficult.[3]The development of NEC has been strongly linked to dysbiosis of the preterm gut,[4] which is thought to promote uncontrolled inflammation of an immunologically immature, intrinsically hyperreactive intestinal epithelium culminating in epithelial and transmural necrosis.[3,5] In support of this theory, proliferation of *Proteobacterial* genera is known to accentuate enteric inflammation and precede NEC.[3,6–8] Moreover, signaling of the intestinal immune receptor for lipopolysaccharide (TLR4: Toll-like Receptor 4) by gram-negative bacteria is necessary for the development of NEC in animal models and, conversely, NEC cannot be induced in germ-free animals.[5,9]

Changing the composition of the intestinal microbiota through enteral probiotic supplementation may promote a microbial community that attenuates or even prevents dysbiotic NEC. Meta-analyses of studies of probiotic supplementation in more than 10,000 patients indicate with varying degrees of certainty that probiotics can reduce rates of NEC and associated mortality in preterm infants.[10–12] The generalizability of these results is limited by negative trials and by a high degree of heterogeneity in probiotic formulation. As a result, it remains controversial whether to use probiotics to prevent NEC due to dysbiosis, and what strain formulation, timing, dosing and feeding regimen to employ.

*Bifidobacterium longum* subspecies *infantis* (*B. infantis*) is a mutualist colonizer of the human infant gut worldwide.[13,14] the strain *B. infantis* EVC001 encodes the complete gene cluster needed to metabolize the full range of prebiotic human milk oligosaccharides (HMOs), complex sugars in human milk that are otherwise indigestible by the infant.[15,16] In turn, *B. infantis* EVC001 functions as a natural symbiont in the infant gut, conferring ecosystem services such as colonization resistance to pathogens and bioactive metabolite production.[13,17–19]

Prior research has demonstrated that supplementing *B. infantis* EVC001 to human milk-fed preterm infants in a neonatal intensive care unit (NICU) is safe, well-tolerated, colonizes the intestinal microbiota, lowers the abundance of NEC-associated pathogenic bacteria and fecal antibiotic resistance genes, and decreases enteric inflammation as measured by fecal calprotectin and cytokine levels.[7] Notwithstanding these promising strain-specific findings, *B. infantis* EVC001 as a *single-strain* probiotic has yet to be associated with a reduction in NEC in preterm infants.[20,21] Accordingly, the objective of this study was to evaluate the impact of *B. infantis* EVC001 supplementation on rates of NEC in at-risk preterm infants.

## Methods

### Study design

A non-concurrent, retrospective cohort design was used to compare clinical outcomes in VLBW infants supplemented with *B. infantis* EVC001 (Evivo®, Evolve BioSystems, Davis, CA) from initiation of trophic feeding until 34 weeks post-menstrual age (PMA) or two weeks, whichever was greater, with VLBW infants who did not receive *B. infantis* EVC001. Study approval was granted by the Oregon Health & Science University (OHSU) Institutional Review Board (IRB #20336).

### Data source

Data were collected from electronic medical record review of infants admitted to the Level IV NICU at Doernbecher Children’s Hospital of OHSU from January 2014 to November 2020. Data were identified using both the OHSU Epic Research Data Warehouse correlated with the OHSU Vermont Oxford Network data until complete correlation was achieved. A minimum of two reviewers independently validated collected data.

### Eligibility criteria

Eligible infants (1) weighed less than 1500 grams at birth, (2) received full resuscitation and survived until day-of-life 3 (the earliest time at which VLBWs would have received at least one feed of EVC001), (3) were fed human milk-based diets consisting of either mother’s milk, donor milk or a combination thereof, (4) were fed according to institutional guidelines incorporating best practices for NEC prevention, including a human milk-based diet, an initial period of trophic feeding and gradual feeding advancements, and (5) did not have hemodynamically significant congenital heart disease.

### Exclusion criteria

Excluded infants (1) underwent palliative delivery or unsuccessful resuscitation, (2) died prior to day-of-life 4, (3) were fed a non-human milk-based diet prior to 34 weeks PMA, (4) had immunodeficiency, or (5) if in the EVC001 group, received less than two feeds of EVC001.

### Cohorts

The reference, unexposed cohort (No EVC001) was defined as VLBW infants admitted to the NICU between January 2014 and May 2018 who were not supplemented with *B. infantis* EVC001. The exposed cohort (EVC001-fed) was defined as VLBW infants admitted to the NICU between June 2018 and November 2020 who received at least two doses of *B. infantis* EVC001. Inclusion criteria for the EVC001-fed cohort required that infants receive two or more doses of B. infantis EVC001 to ensure intestinal colonization..

### Core human milk-based diet

Details are available in Supplemental Data S1. Briefly, each cohort was fed a human milk-based diet of mother’s milk, donor milk, or both. Feeding regimens consisted of an initial period of trophic feeding followed by daily advancements as tolerated to a goal feeding volume of 150-160 mL/kg/day. Donor milk-feeding was continued until at least 34 weeks PMA or for a minimum of five days if greater than 34 weeks at birth, after which donor milk was replaced with bovine milk-based formula if mother’s milk was unavailable. A bovine milk-based human milk fortifier (Similac® HMF, Abbott, IL) was used to meet the nutrient and energy needs of VLBW infants until September 2017. Thereafter, human milk-based fortification (Prolacta®, Monrovia, CA) was used for ELBW infants as best practice for NEC prevention. Bovine milk-based fortification continued to be used for infants weighing more than 1000 grams but less than 1500 grams at birth. As of March 2020, the use of human milk-based fortification was expanded to all infants with birth weights less than 1250 grams.

### B. infantis EVC001 supplementation

8 billion colony forming units (CFU) of activated *B. infantis* EVC001 suspended in 0.5 mL of medium chain triglyceride oil was supplemented daily to infants in the EVC001-fed cohort via gastric tube prior to a morning feed. From June 2018 to July 2019, *B. infantis* EVC001 supplementation was initiated at feeding volumes of 80-100 mL/kg/day feeding. In August 2019, the supplementation protocol was revised to begin on the second day of trophic feeding. Supplementation was continued until 34 weeks PMA or for a minimum of two weeks, whichever duration was longer.

### Covariates

Variables examined as potential predictors, confounders, and effect-modifiers were birth weight, sex, gestational age at birth, small for gestational age (SGA), the presence of congenital heart disease (CHD), antenatal steroid administration prior to delivery (ANS), defined as one or more maternal doses of betamethasone within two weeks of delivery, mode of delivery and packed red blood cell (PRBC) transfusion within 72 hours of NEC diagnosis. The primary outcome was NEC Bell Stage 2 or greater as abstracted from electronic medical record review, and near real-time case tracking by at least two or more neonatologists.

### Primary outcome measure

The diagnosis of NEC was determined using the modified Bell staging system.[22] NEC was assigned if the infant’s condition met criteria for modified Bell stage 2 or greater. Cases of spontaneous intestinal perforation (SIP) were excluded. SIP was defined as gastrointestinal perforation without signs of NEC. The diagnosis of NEC was confirmed by independent review of each case by neonatologists and pediatric surgeons.

### Secondary outcome measures

Secondary outcome measures were NEC requiring antibiotic therapy and supportive care (medical NEC), NEC requiring operative intervention (surgical NEC), NEC-associated mortality, and day-of-life (DOL) at NEC diagnosis. Modified Bell staging was determined by electronic medical record review and real-time case tracking, as described above.

### Statistical analyses

Wilcoxon Rank Sum tests and Pearson’s χ^2^or Fisher’s exact tests were used to determine differences in demographic and clinical characteristics between exposed and unexposed cohorts for continuous and discrete variables. Log-binomial regression models were used to compare risks of NEC-associated outcomes between exposed and unexposed cohorts accounting for potential effect-modifiers and confounding by covariates including sex, birth weight, gestational age at birth and mode of delivery. Risk ratios and adjusted number needed to treat were calculated as measures of association and exposure effect. Moderation effects were examined using statistical significances of pairwise interaction terms involving exposure and other covariates. Control for potential confounders was based on fitting models of each confounder one at a time to explore association with NEC. Subgroup analysis of effects of Prolacta on the No EVC001 cohort was performed on infants < 1000g at birth. Additionally, analysis of NEC rates between cohorts was performed, separately, on infants < 1000g at birth and ≥ 1000g at birth.

Overall NEC rates with 95% confidence intervals (CI) were calculated for exposed and unexposed cohorts for VLBW infants of all birth weights and for a subgroup of ELBW infants. Rates of NEC-associated mortality, medical NEC and surgical NEC in both cohorts were compared by Fisher’s exact test. Confidence intervals were calculated using a normal approximation and, for data with few outcomes, a continuity correction was applied.

Relative risk of NEC by cohort was estimated with a log-binomial regression model adjusting for sex, birth weight, gestational age at birth and mode of delivery. Adjusted number needed to treat was calculated from the cohort coefficient of the model. The same model and calculations were applied to a subgroup of ELBW infants.

Statistical significance was assessed at an alpha of 0.05. All statistics were performed with R version 3.6.3 with the ‘tidyverse’, ‘ggpubr’, ‘psych’, ‘kableExtra’, ‘car’, ‘e1071’, ‘epitools’, ‘emmeans’, ‘meta’, ‘cmprsk’ and ‘survival’ packages by collaborating biostatisticians at Indiana University School of Public Health.

## Results

588 VLBW infants were identified during the study period. Of these, 105 infants were palliatively delivered, unsuccessfully resuscitated, died within the first 3 days of life, or not fed human milk-based diets (Figure 1). 483 remaining infants met inclusion criteria and consisted of 301 infants who were not exposed to *B. infantis* EVC001 (No EVC001) and 182 infants who were exposed to the probiotic (EVC001-fed). There were no significant differences in measured covariates between the two cohorts with the exception of sex and antenatal steroid administration (Table 1). The mean gestational age at birth for both cohorts was 28 weeks, with a mean birth weight for the No EVC001 cohort of 1045.4 grams (range: 325-1490 grams) and 1048.0 grams (range: 358-1498 grams) for the EVC001-fed cohort. The majority of infants were delivered by Caesarian section. There was a statistically significant lower percentage of antenatal steroid administration in the EVC001-fed cohorts (83.5% EVC001-fed vs. 90.4% No EVC001, P = 0.0259), and female infants (42.3% EVC001-fed vs 52.2% No EVC001, P = 0.0358). There were no significant differences between cohorts for any of the measured covariates in the subgroup analysis of ELBW infants.

**Figure 1:**
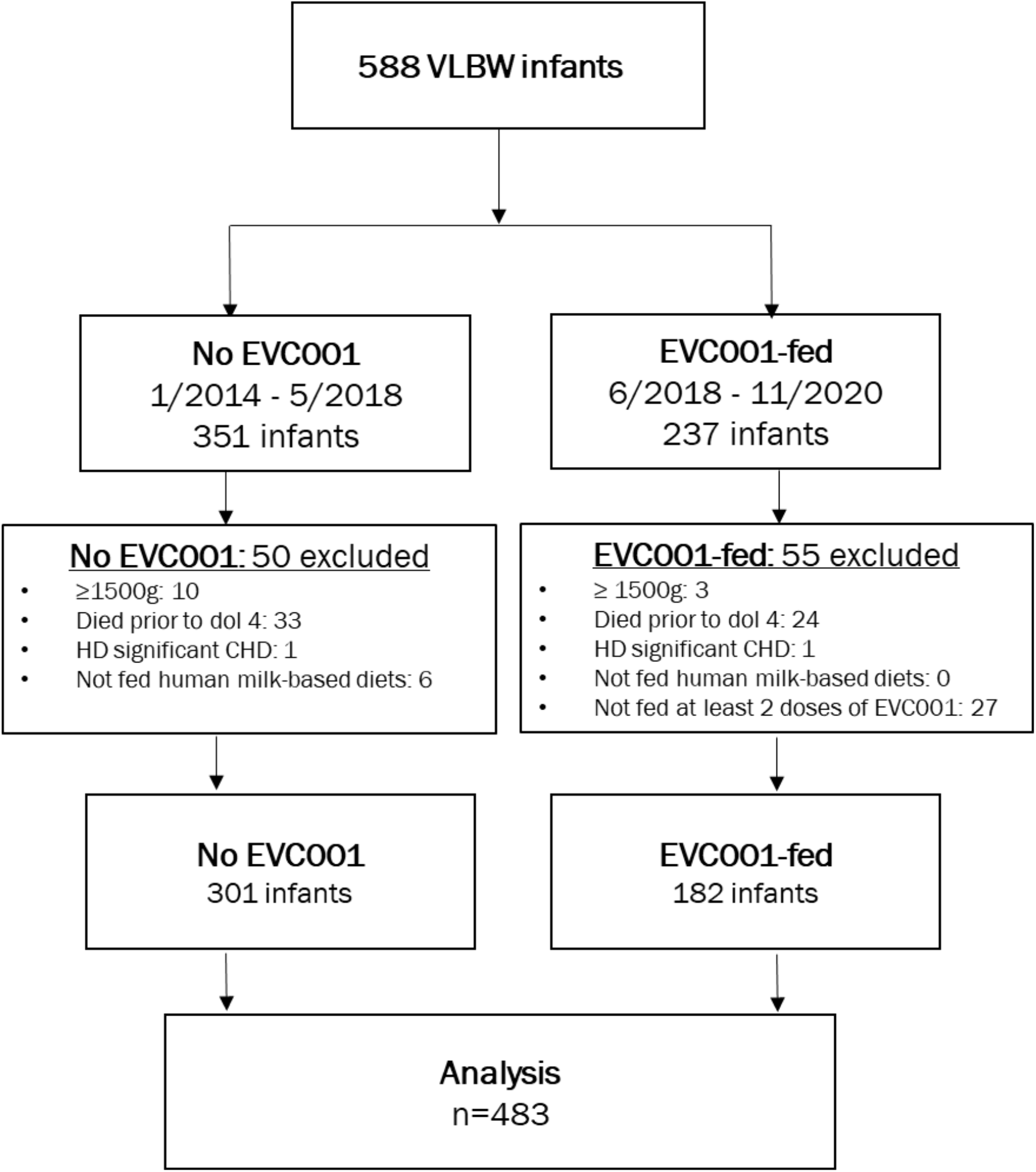
STROBE flow diagram identifying infants excluded from analysis.

**Table 1:**
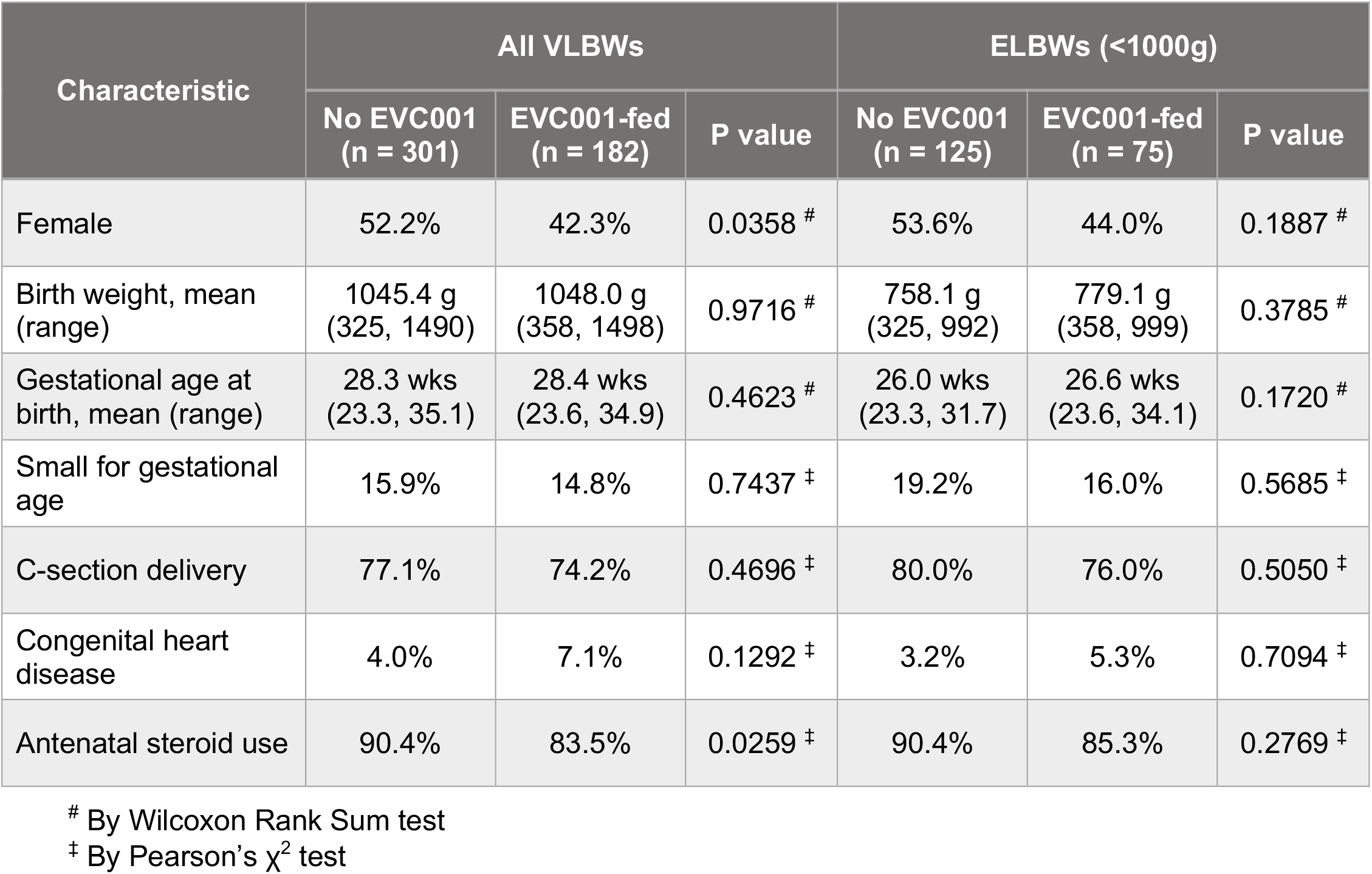
Demographics.

| Characteristic | All VLBWs |  |  | ELBW (<1000g) |  |  |
| --- | --- | --- | --- | --- | --- | --- |
|  | No EVC001<br>(n = 301) | EVC001-fed<br>(n = 182) | P value | No EVC001<br>(n = 125) | EVC001-fed<br>(n = 75) | P value |
| Female | 52.2% | 42.3% | 0.0358 <sup>#</sup> | 53.6% | 44.0% | 0.1887 <sup>#</sup> |
| Birth weight, mean (range) | 1045.4 g<br>(325, 1490) | 1048.0 g<br>(358, 1498) | 0.9716 <sup>#</sup> | 758.1 g<br>(325, 992) | 779.1 g<br>(358, 999) | 0.3785 <sup>#</sup> |
| Gestational age at birth, mean (range) | 28.3 wks<br>(23.3, 35.1) | 28.4 wks<br>(23.6, 34.9) | 0.4623 <sup>#</sup> | 26.0 wks<br>(23.3, 31.7) | 26.6 wks<br>(23.6, 34.1) | 0.1720 <sup>#</sup> |
| Small for gestational age | 15.9% | 14.8% | 0.7437 <sup>‡</sup> | 19.2% | 16.0% | 0.5685 <sup>‡</sup> |
| C-section delivery | 77.1% | 74.2% | 0.4696 <sup>‡</sup> | 80.0% | 76.0% | 0.5050 <sup>‡</sup> |
| Congenital heart disease | 4.0% | 7.1% | 0.1292 <sup>‡</sup> | 3.2% | 5.3% | 0.7094 <sup>‡</sup> |
| Antenatal steroid use | 90.4% | 83.5% | 0.0259 <sup>‡</sup> | 90.4% | 85.3% | 0.2769 <sup>‡</sup> |

<sup>#</sup> By Wilcoxon Rank Sum test<sup>‡</sup> By Pearson's $\chi^2$ test

With regard to the primary outcome, there were 33 cases of NEC (11.0%) in the No EVC001 cohort (Figure 2, Figure 3, Table 2), inclusive of the nine-month period after introduction of human milk-based fortification in the ELBW population. No significant difference was observed in rates of NEC between ELBW infants in the No EVC001 cohort during this nine-month period of exclusive human milk use [19 of 95 ELBWs (20.0%) pre-human milk-based fortification compared to 5 of 30 ELBWs (16.7%) post-human milk-based fortification, P = 0.795]. Due to similar NEC rates before and after initiation of human milk-based fortification, the No EVC001 cohort was analyzed as a single group for all outcomes, regardless of human milk fortification type.

**Figure 2:**
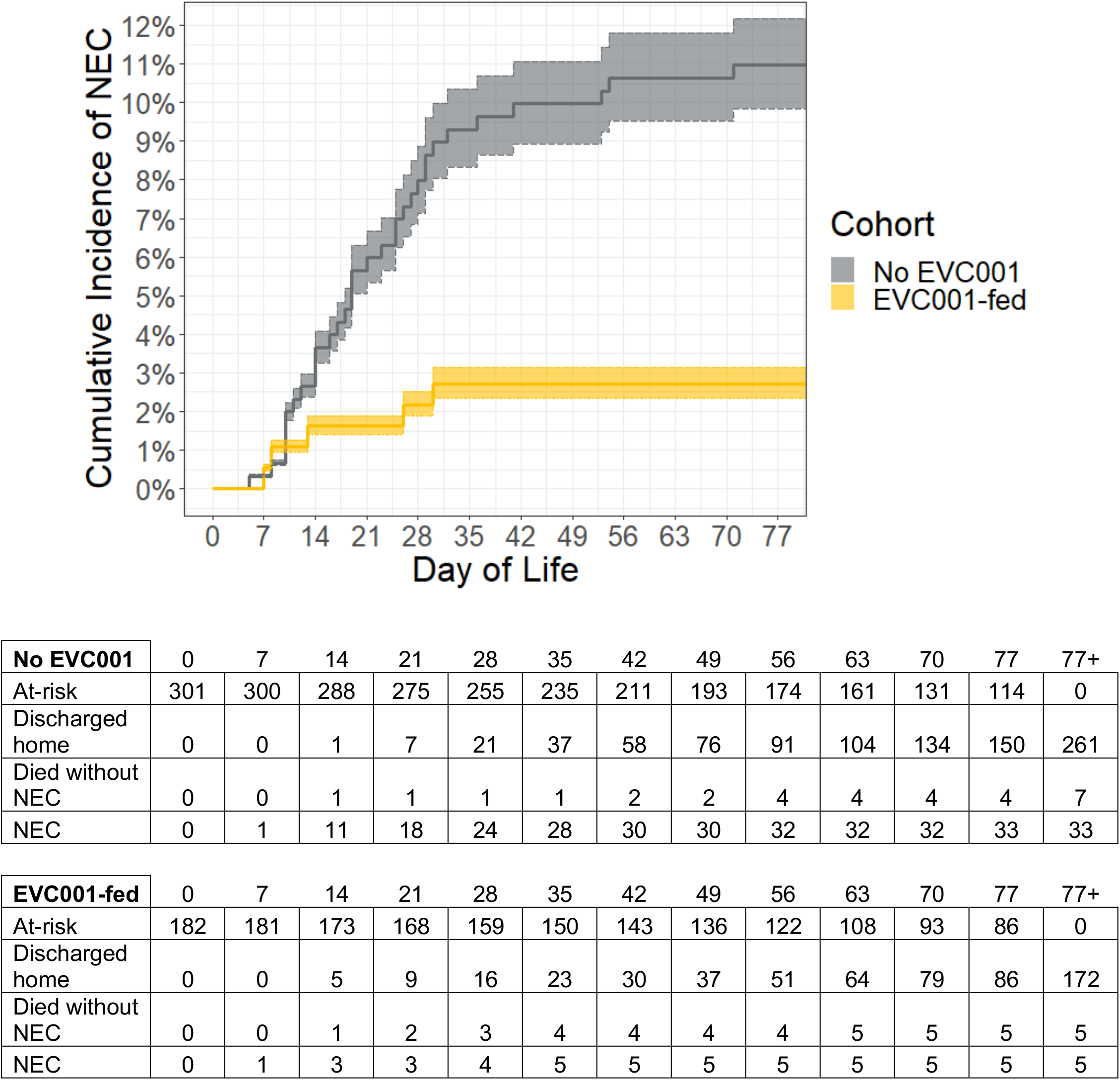
Cumulative incidence of NEC by day-of-life. Shaded regions show the 95% confidence intervals around the estimates. Table indicates total number of infants discharged home, infants who died without NEC, and infants who developed NEC, weekly.

**Figure 3:**
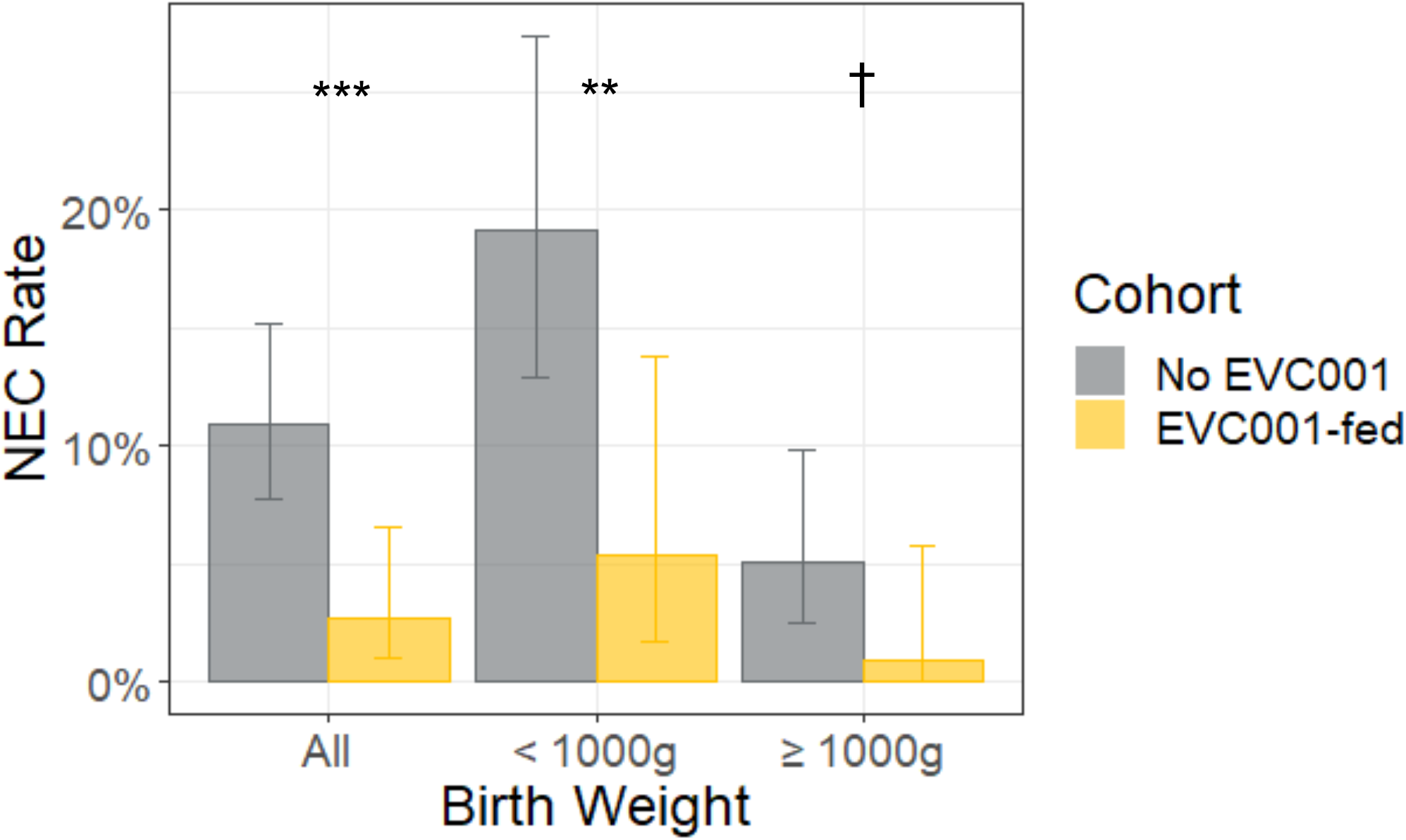
NEC rates by birth weight and cohort. Error bars show the 95% confidence intervals around the estimates. Results from Fisher’s exact test are shown as ***(P < 0.001), **(P < 0.01), and †(P < 0.1).

**Table 2:** NEC Outcomes.

| Outcome | All VLBWs |  |  | ELBW (<1000 g) |  |  |
| --- | --- | --- | --- | --- | --- | --- |
|  | No EVC001<br>(n = 301) | EVC001-fed<br>(n = 182) | P value | No EVC001<br>(n = 125) | EVC001-fed<br>(n = 75) | P value |
| NEC | 11.0% | 2.7% | 0.0008 ‡ | 19.2% | 5.3% | 0.0059 ‡ |
| Risk ratio<br>(95% C.I.) | 0.270<br>(0.094, 0.614) | | 0.0054 \$ | 0.283<br>(0.085, 0.698) | | 0.0167 \$ |
| Number needed<br>to treat<br>(95% C.I.) | 12.4<br>(10.0, 23.5) |  |  | 7.3<br>(5.7, 17.3) |  |  |
| NEC-related<br>mortality | 2.7% | 0% | 0.0274 ‡ | 5.6% | 0% | 0.0468 ‡ |
| NEC onset, mean<br>day of life | 23.5 | 16.8 | 0.2994 # | 24.0 | 19.3 | 0.6453 # |
| Medical NEC<br>(Bell Stage II) | 7.0% | 1.1% | 0.0032 ‡ | 12.0% | 2.7% | 0.0332 ‡ |
| Surgical NEC<br>(Bell Stage III) | 4.0% | 1.6% | 0.1836 ‡ | 7.2% | 2.7% | 0.2147 ‡ |

‡ By Fisher's exact test
\$ By log binomial regression adjusted for sex, birth weight, gestational age at birth and mode of delivery

There were 5 cases of NEC (2.7%) in the EVC001-fed cohort. The difference in rates of NEC between cohorts was statistically significant (P = 0.0008). Log-binomial models demonstrated that infants in the EVC001-fed cohort had a 73% risk reduction of NEC compared to infants in the No EVC001 cohort (adjusted risk ratio 0.270, 95% CI 0.094, 0.614, P = 0.0054) after adjusting for differences in sex, birth weight, gestational age and mode of delivery. The adjusted number needed to treat based on these outcomes was 12.4 (95% CI 10.0, 23.5). There were 6 additional incidences of NEC during the EVC001-fed epoch in VLBW infants without hemodynamically significant congenital heart disease who received no (n=5) or one (n=1) feeding of EVC001 prior to their diagnosis of NEC.

Subgroup analysis was carried out to determine the effect of *B. infantis* EVC001 supplementation on ELBW infants with a birth weight less than 1000 grams and infants with a birth weight between 1000 and 1499 grams. ELBW infants demonstrated a statistically significant difference in NEC rates between cohorts (No EVC001 n = 125, EVC001-fed n = 75, 19.2% versus 5.3%, P = 0.0059) with an adjusted risk ratio among ELBW infants of 0.283 (95% CI 0.085, 0.698, P = 0.0167) (Figure 3, Table 2). The adjusted number needed to treat was 7.3 (95% CI 5.7, 17.3). Of note, ELBW infants in the EVC001-fed cohort received human milk-based fortification whereas ELBW infants in the No EVC001 cohort received bovine milk-based fortification until the final nine months of that cohort period (n = 30). Infants with a birth weight between 1000 and 1499 grams also showed a decrease in NEC rates between the No EVC001 (9/176; 5.1%) and EVC001-fed cohorts (1/107; 0.9%) that had a trend towards significance (P = 0.0955).

With regard to secondary outcomes, when evaluating the severity of NEC, rates of both medical NEC and surgical NEC decreased in the EVC001-fed cohort. There was a statistically significant difference in the rates of medical NEC [7.0% (n = 21) versus 1.1% (n = 2), P = 0.0032] and a trend toward a difference in the rates of surgical NEC [4.0% (n = 12) versus 1.6% (n = 3), P = 0.1836] (Figure 4). Among infants who developed NEC, there was no statistically significant difference between rates of medical versus surgical NEC (P = 0.3649).

**Figure 4:**
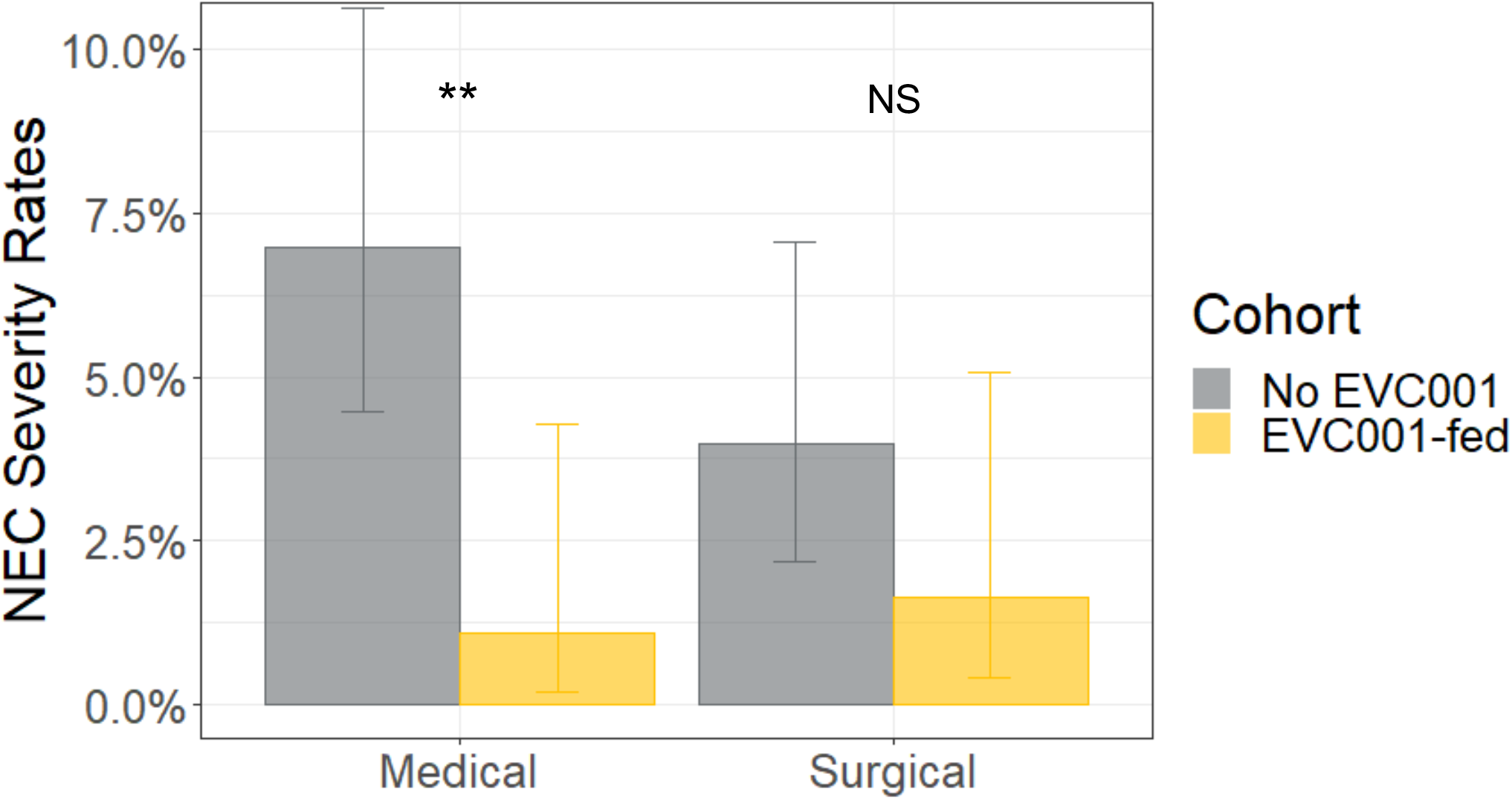
NEC rates by severity (medical and surgical). Error bars show 95% confidence intervals around the estimates. Results from Fisher’s exact test are shown as **(P < 0.01) and NS (P > 0.1).

There was zero NEC-associated mortality in infants supplemented with *B. infantis* EVC001 compared to a NEC-associated mortality of 2.7% of all infants in the No EVC001 cohort (Figure 5, Table 2). The difference in NEC-associated mortality between cohorts was statistically significant (P = 0.0274). In the ELBW subgroup analysis of NEC-associated mortality, there was also a statistically significant difference [5.6% (n = 7) in No EVC001 cohort versus 0% (n = 0) in EVC001-fed cohort, P = 0.0468]. For VLBW infants diagnosed with NEC, while the difference in NEC mortality, 24.2% (8 of 33) for infants diagnosed with NEC in the No EVC001 cohort compared to 0% (0 of 5) in the EVC001 fed cohort, was notable, it was not statistically significant (p=0.548). Likewise, the mean day of life of onset of NEC for VLBWs (23.5 days for the No EVC001 cohort versus 16.8 days for the EVC001 fed cohort) was not significant (p=0.2995) (Table 5). The same was true for mean day of onset of NEC in the ELBW subgroup.

**Figure 5:**
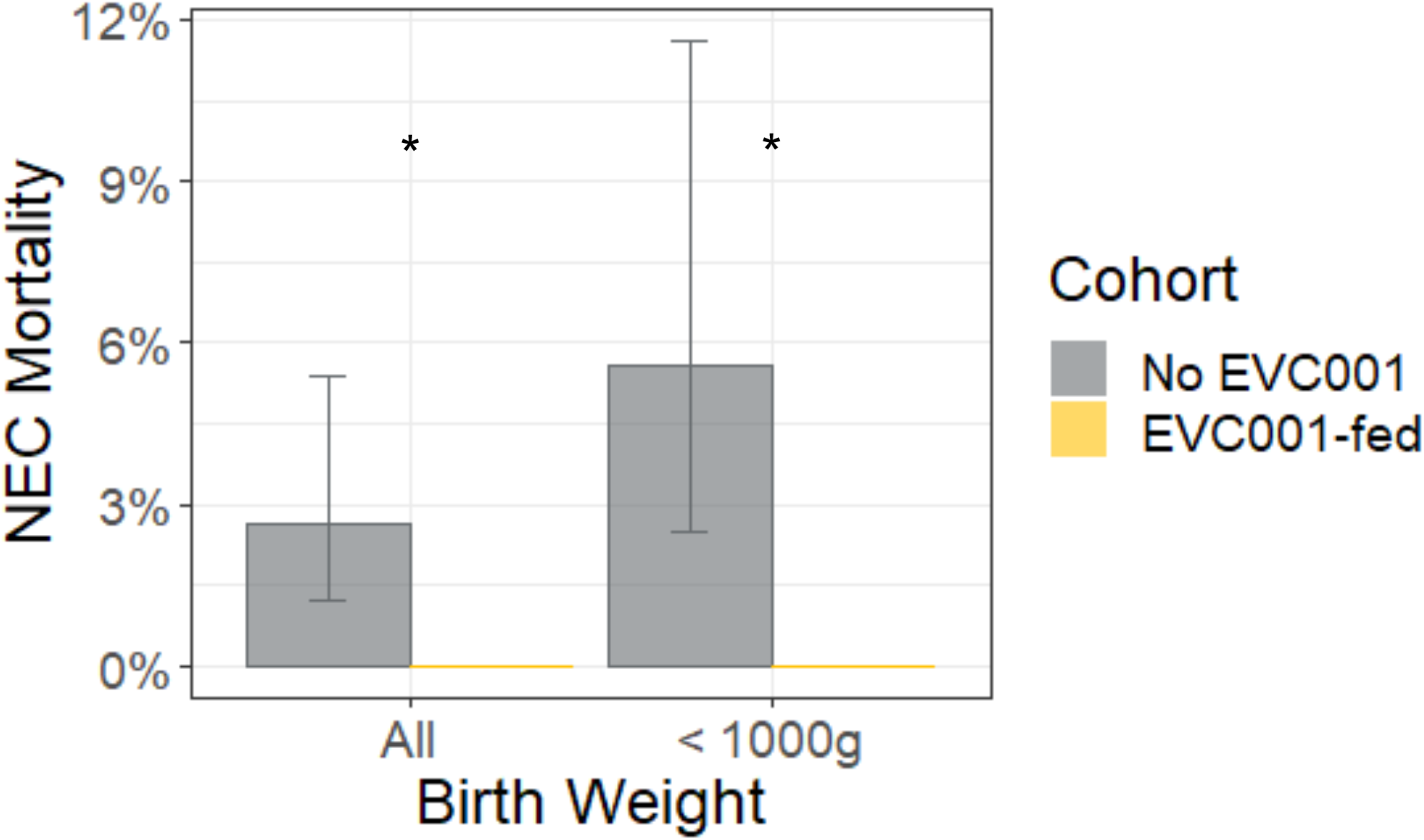
NEC-related mortality rates by birth weight and cohort. Error bars show 95% confidence intervals around the estimates. Result from Fisher’s exact test is shown as *(P < 0.05).

Finally, no adverse effects were observed with *B. infantis* EVC001 supplementation, including no cases of *B*. infantis bacteremia or other infection.

## Discussion

In this retrospective cohort study of 483 VLBW infants in a single center Level IV NICU over nearly seven years, we observed a significant reduction in necrotizing enterocolitis among VLBW human milk-fed infants associated with daily administration of the infant gut symbiont *B. infantis* EVC001. The NEC rate in those infants given more than one feeding of EVC001 decreased from 11% to 2.7%, a 73% risk reduction. There were no cases of NEC-associated mortality in the EVC001-fed cohort, which was significant for both the overall EVC001-fed cohort and the ELBW subgroup. By comparison, 24.2% of infants who developed NEC in the No EVC001 cohort died. Importantly, there were no adverse events associated with *B. infantis* EVC001 supplementation, including *B. infantis* bacteremia.

There were care improvement measures that were undertaken during the study period, a small number of which were aimed at optimizing nutrition and NEC reduction. Most relevant was the use of an exclusive human milk diet for ELBW infants, followed by expansion to infants <1250 grams,. Although there were no differences between cohorts of VLBWs with respect to mode of delivery, birth weight, gestational age at birth, small for gestational age and the presence of congenital heart disease, there were marginally significant differences in rates of antenatal steroid administration and sex between cohorts. Given that antenatal steroids reduce the risk for NEC in preterm infants,[23] a higher rate of antenatal steroid use in the No EVC001 cohort would have been more likely to mitigate the observed benefit of *B. infantis* EVC001 supplementation. Similarly, a lower proportion of females in the EVC001-fed cohort might also reduce potential benefits of EVC001 supplementation.[24] Taken together, these findings support the safe use of *B. infantis* EVC001 as a probiotic to reduce the incidence of NEC in the most at-risk NICU patients.

There is mechanistic evidence to support a potential role for *B. infantis* in NEC reduction [25]. *B. infantis* is known to be a foundational colonizer of the human milk-fed infant gut that has co-evolved to optimally metabolize human milk oligosaccharides. *B. infantis* EVC001 harbors genes that facilitate the intracellular transport and metabolism of the full array of HMOs,[26] properties that have thus far not been identified in other *B. infantis* strains or other bacterial species.[16,27] Oligosaccharide transporters and glycosidases confer a growth advantage, which enables *B. infantis* EVC001 to predominate within the human milk-fed infant gut.

In turn, *B. infantis* EVC001 colonization is hypothesized to provide important ecosystem services.[18] *B. infantis* EVC001 confers colonization resistance to pathogens by competitive growth and by fermenting HMOs into organic acids such as lactate and acetate that reduce intestinal pH and inhibit the growth of pathogenic species, including *Enterobacteriaceae* and *Clostridia*.[28] Fermentation products also strengthen intestinal barrier function and exert anti-inflammatory effects.[7,29,30] In particular, indole-containing tryptophan metabolites, enriched in the stools of human milk-fed infants supplemented with *B. infantis* EVC001, have been shown to down-regulate TLR4 signaling via the aryl hydrocarbon receptor pathway.[31–33]

Human milk feeding in concert with this evolutionarily co-evolved bacterium may therefore influence the intestinal microbiota through colonization resistance, modulation of the intestinal inflammatory environment, and the host immune system. Consistent with this concept, *B. infantis* EVC001 has been shown to reduce the burden of antibiotic resistance genes in the stool of human milk-fed term and preterm infants,[7,34] reduce fecal markers of intestinal inflammation,[7] and modulate systemic inflammation,[25] possibly in a sustained manner.

This is the first report of an association between *B. infantis* EVC001 and VLBW NEC risk reduction. Of 56 trials included in the most recent Cochrane meta-analysis (2020) of the use of probiotics to prevent NEC, 14 trials used single-strain formulations containing other *Bifidobacterium* species; none used *B. infantis*. Trials that included *B. infantis* did so only in multi-strain formulations.[11] Similarly, of 63 trials analyzed in a 2020 American Gastroenterological Association network meta-analysis (spanning research from 1986 to 2019), 15 trials used *B. infantis* in multi-strain formulations, most often combined with *Lactobacillus*, and no studies used *B. infantis* as a single-strain probiotic.[12] Notably, the most effective multi-strain formulations in both meta-analyses contained *B. infantis*.

This report adds to the diverse body of evidence that probiotic use is and effective means of NEC reduction, as well as the challenge to neonatology to provide improved clarity regarding, but not limited to, optimal strain(s), formulation, dosing, duration, target population, and dietary substrate, in particular, an exclusive human milk diet. This includes mechanistic research to provide the biological basis for the reported clinical effects, an area that has been under-resourced in proportion to the impact of NEC. Even less understood are long term effects of microbiome alteration in relation to the developing gut, and overall health. It remains difficult to interpret the large quantity of available evidence to select a single- or multi-strain probiotic formulation for the prevention of NEC in preterm infants. Based on outcomes observed in this study and in light of its unique symbiotic, genetic, ecological, and biochemical properties, *B. infantis* EVC001 represents a promising candidate. More broadly, the results of this study suggest that a given probiotic formulation must have established efficacy, quality, and a viable mechanism of action in the infant gut.

## Limitations

The results of this study are limited by its retrospective, observational design, and by an absence of fecal sampling to confirm that *B. infantis* EVC001 supplementation led to successful modulation of the preterm intestinal microbiota. Thus, the observed reduction in NEC rate associated with *B. infantis* EVC001 is limited to an association and not necessarily causal. During the nearly 7-year chart review period, there were evolutions in patient care and unmeasured confounders that may have influenced the difference in NEC rates, including the addition of human milk-based fortifier for ELBW infants during the late No EVC001 period, followed by the addition to 1000-1249 gram BW infants late in the EVC001 exposed period. While the addition of an exclusive human milk diet for ELBW infants did not result in a detectable decrease in NEC in the No EVC001 epoch, the time period was insufficient to detect a potential effect on NEC reduction in this population. The combination, however, of an exclusive human milk diet and EVC001 yielded an apparent reduction in NEC that remains important. Other care changes implemented during the period applied to both epochs, did not apply to care thought to affect NEC, and therefore would not be anticipated to have a large impact on the observed data. Finally, while there was an absence of fecal colonization data from EVC001 recipients, this has been well demonstrated in recent publications.[7,17] .

## Conclusion

In this retrospective, electronic medical record review, administration of *Bifidobacterium longum* subspecies *infantis* EVC001 as a single strain to VLBW infants is associated with a significant decrease in NEC and NEC-associated mortality, including ELBW infants. The effect among ELBW infants was observed in combination with an exclusive human milk diet. The NEC reduction was free of any associated adverse events. Probiotic supplementation in this study was part of a comprehensive NEC prevention strategy implemented for all infants less than 1500 grams at birth. Based on these findings, *B. infantis* EVC001 supplementation can be considered a safe and effective strategy for prevention of NEC in the NICU and for modifying dysbiosis thought to underpin a significant proportion of NEC.

## Supporting information

Strobe Checklist

Conflict of Interest form for Brian Scottoline

## Data Availability

Data will be made available upon acceptance for journal publication.

## Abbreviations

ANS: Antenatal steroid
ELBW: Extremely low birth weight
HMF: Human milk fortifier
HMOs: Human milk oligosaccharides NEC Necrotizing enterocolitis
NICU: Neonatal intensive care unit
OHSU: Oregon Health & Science University PMA Post-menstrual age
VLBW: Very low birth weight

